# Pulmonary function and comparative SARS-CoV-2 RBD-specific IgG antibody response among the COVID-19 recovered group

**DOI:** 10.1101/2025.02.17.25321314

**Authors:** Abu Bakar Siddik, Arman Faisal, Abdullah-Hel-Kafi khan, Md. Meer Mahbubul Alam, Jannatul Nayeem, Umme Kulsum, Sharmin Aktar Mukta, Zannat Kawser, Imrul Hasan, Kasrina Azad, Moajjam Hossain, Sanchita Kar, Nishat Sultana, Md Rabiul Alam, Ahmed Mustafa, Mohammad Tanbir Habib, Edward T. Ryan, Firdausi Qadri, Mohammad Rashidul Hassan

**Affiliations:** *Infectious Disease Division, institute for developing Science and Health initiatives, Dhaka, 1206, Bangladesh; Department of Respiratory Medicine, Ingenious Health Care Limited, Dhaka, 1207, Bangladesh; Infectious Disease Division, international center for diarrhoeal disease research (icddr,b), Dhaka, 1212, Bangladesh; Department of Biochemistry and Molecular Biology, University of Dhaka, Dhaka, 1000, Bangladesh; Department of Immunology and Infectious diseases, Harvard T.H. Chan School of Public Health, Boston, Massachusetts, MA 02115, USA; Department of Medicine, Harvard Medical School, Boston, Massachusetts, MA 02115, USA; Anwar Khan Medical College, Dhaka, 1207, Bangladesh

**Keywords:** COVID-19, SARS-CoV-2, Pulmonary Function, Long-COVID, Antibody response

## Abstract

Coronavirus disease 2019 (COVID-19) is a highly contagious infectious disease caused by severe acute respiratory syndrome coronavirus 2 (SARS-CoV-2) responsible for millions of deaths and substantial morbidity worldwide. Several studies report that up to 50% of individuals who recover from acute SARS-CoV-2 infection experienced a plethora of long-COVID symptoms for weeks, months, or even up to a year. Abnormal pulmonary function is one of the most critical manifestations of long-COVID even after recovering from COVID-19. Understanding the long-term pulmonary consequences and immune response among individuals recovering from COVID-19, who experienced disease severity ranging from mild to severe symptoms, is crucial for comprehensive post-recovery care and vaccination strategies.

This prospective case-control study included 29 individuals who had recovered from COVID-19 with a history of mild to severe symptoms and 64 controls. Assessments of pulmonary functional measures, such as FVC, FEV1, FEV1/FVC ratio, FEF, MEF, and PEF were carried out following recovery from COVID-19. Additionally, IgG antibody responses were examined by ELISA for up to six months through multiple follow-ups following two doses of vaccination, with an additional follow-up 30 days after the booster dose (third dose).

Pulmonary functional abnormalities were prevalent in the recovered group who previously exhibited different symptoms (53% mild, 66% moderate, and 50% severe) compared to the control group (23%). Higher IgG antibody titers were observed among the recovered groups, significantly elevated in severe and moderate cases following vaccination. Following vaccination, the recovered group who experienced disease history showed significantly higher titers at day 14, particularly in severe (1418 IU/mL) and moderate (1390 IU/mL) groups compared to the control group (968 IU/mL) (p<0.005). Notably, antibody titers are negatively correlated with pulmonary function test (PFT) parameters such as Forced Vital Capacity (FVC) and Forced Expiratory Volume in 1 second (FEV1). All groups experienced a significant (p<0.005) decrease in antibody titers within 90-120 days of two doses of vaccination. After five to six months, the titers were comparable to the baseline titer, indicating the importance of vaccination and booster doses regardless of previous infection history. Overall, our study underscores the significance of pulmonary function assessment post-COVID-19 recovery for long-term respiratory health and emphasizes the importance of vaccination regardless of infectious history. To ensure long-term respiratory health, this study emphasizes the significance of evaluating pulmonary function in those who might have contained asymptomatic COVID-19 infections as well as those who have recovered from symptomatic COVID-19 infections. Furthermore, these findings underscore the importance of vaccination regardless of infection history as a key strategy in pandemic preparedness. To assess the impact of long-COVID on respiratory health, this study underscores the importance of evaluating pulmonary function in individuals, whether they had symptomatic or asymptomatic COVID-19. Furthermore, the findings from the immune response analysis highlight the critical role of vaccination, regardless of infection history, as a key strategy of pandemic preparedness.

## 2. Introduction

Severe Acute Respiratory Syndrome Coronavirus 2 (SARS-CoV-2) causes Coronavirus Disease 2019 (COVID-19), a highly transmissible viral infection that has had a profound impact on global public health. The World Health Organization (WHO) declared COVID-19 a pandemic in March 2020, and it has since been responsible for over 600 million cases worldwide, with significant mortality and morbidity (1,2). In Bangladesh, the first COVID-19 case was identified on March 8, 2020 (3).

Patients with COVID-19 may exhibit a variety of subclinical and clinical manifestations, ranging from mild infection to severe respiratory disorders, systemic disease, and multiorgan failure (4).

Infection with SARS-CoV-2 typically affects the lung, where patients with COVID-19 have diffused alveolar damage, bronchiolitis, alveolitis, and interstitial fibrosis in their lung pathology (5,6). Even after recovering from COVID-19, patients may continue to experience lung issues, which can cause breathing difficulties and result in abnormal results on pulmonary function tests (7). Clinical studies have highlighted that COVID-19 survivors may exhibit persistent lung abnormalities even months after recovery, including diffused alveolar damage, bronchiolitis, and interstitial fibrosis (8,9).

Moreover, in COVID-19 patients with mild to moderate symptoms, spirometry and other pulmonary function tests (PFT) tests showed decreased Forced Vital Capacity (FVC) and total lung capacity (TLC), and symptoms include reduced diffusing capacity, lower exercise capacity, and impaired respiratory-muscle strength (10,11). This poses significant risks not only for individual patients but also for public health, as lingering respiratory issues may increase healthcare demand and contribute to long-term disability, particularly in vulnerable populations such as the elderly and those with pre-existing conditions. A follow-up study of 76 COVID-19 survivors in Wuhan Union Hospital for three months after discharge revealed persistent impairment in Forced Expiratory Volume in the First Second (FEV1), FEV1/FVC ratio, and Diffusing Capacity for Carbon Monoxide (DLCO) in about 32.42% of cases (12).

However, pulmonary dysfunction is influenced by the host-pathogen complex’s interaction, local and systemic immune responses to SARS-CoV-2, and viral spread in the respiratory tract (13,14). The immune response against respiratory viruses like SARS-CoV-2 involves quick innate immune mechanisms followed by later adaptive immune mechanisms (15,16). Innate immune mechanisms, which respond quickly, followed by adaptive immune mechanisms involving B and T lymphocytes, which result in antibody and cytokine-mediated responses, provide defense against respiratory viruses like SARS-CoV-2 (17). Pathogen-specific T cell responses are difficult to measure in routine diagnostic settings, whereas B cell-mediated immunity can be more easily monitored through the quantitative measurement of antibody levels using Enzyme Linked Immunosorbent Assay (ELISA) against SARS-CoV-2 spike (S) glycoprotein and its receptor-binding domain (RBD), as well as the nucleocapsid (N) protein, reported as the most immunogenic (18–20).

Three antibody isotypes like IgM, IgG, and IgA are primarily produced in response to respiratory infections, with IgG remaining in the serum for months to suggest comparatively long-lasting protection (21–23). However, it is well known that the humoral immune response to infection can have both beneficial and detrimental effects, as it can serve as a protective mechanism to resolve the infection while also aggravating tissue injury. In SARS, for example, the IgG response can result in fatal acute lung injury by skewing the inflammation-resolving response (24). The global spread of the SARS-CoV-2 pandemic has been altered by the introduction of vaccines against the virus, lowering both viral transmission and disease burden. On February 7, 2021, Bangladesh began administration of COVID-19 vaccinations. However, immunoassays for the quantitative detection of anti-SARS-CoV-2 antibodies are important due to the fact that there are still a number of cases of breakthrough infections even after receiving the full dose of vaccination (25)]. Moreover, while the long-term antibody response in COVID-19 disease has not yet been fully established, evidence indicates a significant decrease in antibody concentrations 3–4 months after the onset of symptoms as well as among the vaccinated people (26,27). It has been questioned whether administering a third dose of the vaccine, also known as the booster dose, is necessary to increase protection against SARS-CoV-2 infection given the decline in circulating antibodies, which could have an impact on public health policy to mitigate the pandemic by directing vaccination strategies. Therefore, knowledge of the duration and breadth of antibody responses after SARS-CoV-2 infection as well as vaccination is vital for understanding the role that antibodies might play in disease clearance and protection from reinfection and/or severe disease and vaccine effectiveness (28). In addition, these antibodies usually demonstrate a sufficient correlation with antiviral immunity, and anti-receptor-binding domain antibody levels correspond to plasma viral neutralizing activity as well as disease severity (29). According to earlier research, SARS-CoV-2 antibody titers are correlated with severe clinical manifestations of COVID-19. Nevertheless, it is still unknown if clinical manifestations cause a high antibody response or if a high SARSCoV2 antibody titer exacerbates clinical manifestations (30,31). Since SARS-CoV-2-specific antibody concentrations in the clinical course of COVID-19 are still debatable, previous infection and pre-existing antibodies may have some bearing on the severity of the disease and vaccine effectiveness.

Considering a social perspective, long-term respiratory impairments can limit an individual’s ability to work, study, and participate in daily activities, leading to economic hardship and social isolation (13). This is particularly concerning in low- and middle-income countries (LMICs) like Bangladesh, where many families rely on manual labor for income, and health-related disabilities can have a ripple effect on entire households (32).

Therefore, understanding the long-term impact of COVID-19 on lung health is critical to designing public health strategies that address the physical, mental, and economic consequences of the pandemic.

This study aims to explore the long-term sequel of SARS-CoV-2 infection on pulmonary health among the COVID-19 recovered group. Specifically, association between disease severity history (ranging from mild to severe) and pulmonary function as well as immune response dynamics following vaccination.

## 3. Materials and Methods

### 3.1 Study population and sites

This was a prospective case-control longitudinal study conducted by the Institute for Developing Science and Health Initiatives (ideSHi), Dhaka in collaboration with Ingenious Health Care Limited, Dhaka, from November 2021 to November 2022 in Bangladesh. The National Research Ethics Committee (NREC) of the Bangladesh Medical Research Council (BMRC) has approved the study (Ref: BMRC/NREC/2019-2022/432; Registration Number: 41318052021). According to the regulations and guidelines of the “Declaration of Helsinki,” written informed consent was obtained from each participant.

Following informed consent, participants underwent RT-PCR testing for SARS-CoV-2 using Nucleic Acid Diagnostic Kit, 2019-nCoV (Sansure Biotech Inc, Changsha, PR China) following the manufacturer’s instructions. Those who tested negative by RT-PCR and aged ≥ 18 years were eligible to enroll in the study. A total of 93 participants were enrolled in this study, of whom 29 recovered from COVID-19 with different disease severity histories and 64 were control. All the participants who recovered from COVID-19 had symptomatic disease his- tory and were further divided into three groups including mild, moderate, and severe following WHO guidelines depending on their prior infection histories such as clinical symptoms and oxygen saturation (SpO2) report (33).

The control group was defined as individuals who never tested positive for SARS-CoV-2 by RT-PCR and never exhibited COVID-19 symptoms at any point during the pandemic before enrollment in this study. Participants were recruited from Anwar Khan Modern Medical College and Hospital, Ingenious Health Care Limited, and the community level in the Dhaka division. After enrollment, sociodemographic and clinical information were collected using a standardized questionnaire developed for this study **(Supplementary file S1)**. All the enrolled participants received two doses of the Pfizer–BioNTech COVID-19 vaccine following enrollment in the study. Further, they received a booster dose six months later, following the Bangladesh government vaccination policy. Blood samples were collected at nine different time points: once before vaccination, followed by seven additional collections after the two vaccine doses on days 14, 30, 60, 90, 120, 150, and 180, and finally, one sample 30 days after the booster dose to comprehensively evaluate the immune response over time. Collected blood specimens were subjected to several laboratory tests over the study period **(Fig 1)**.

**Fig. 1.**
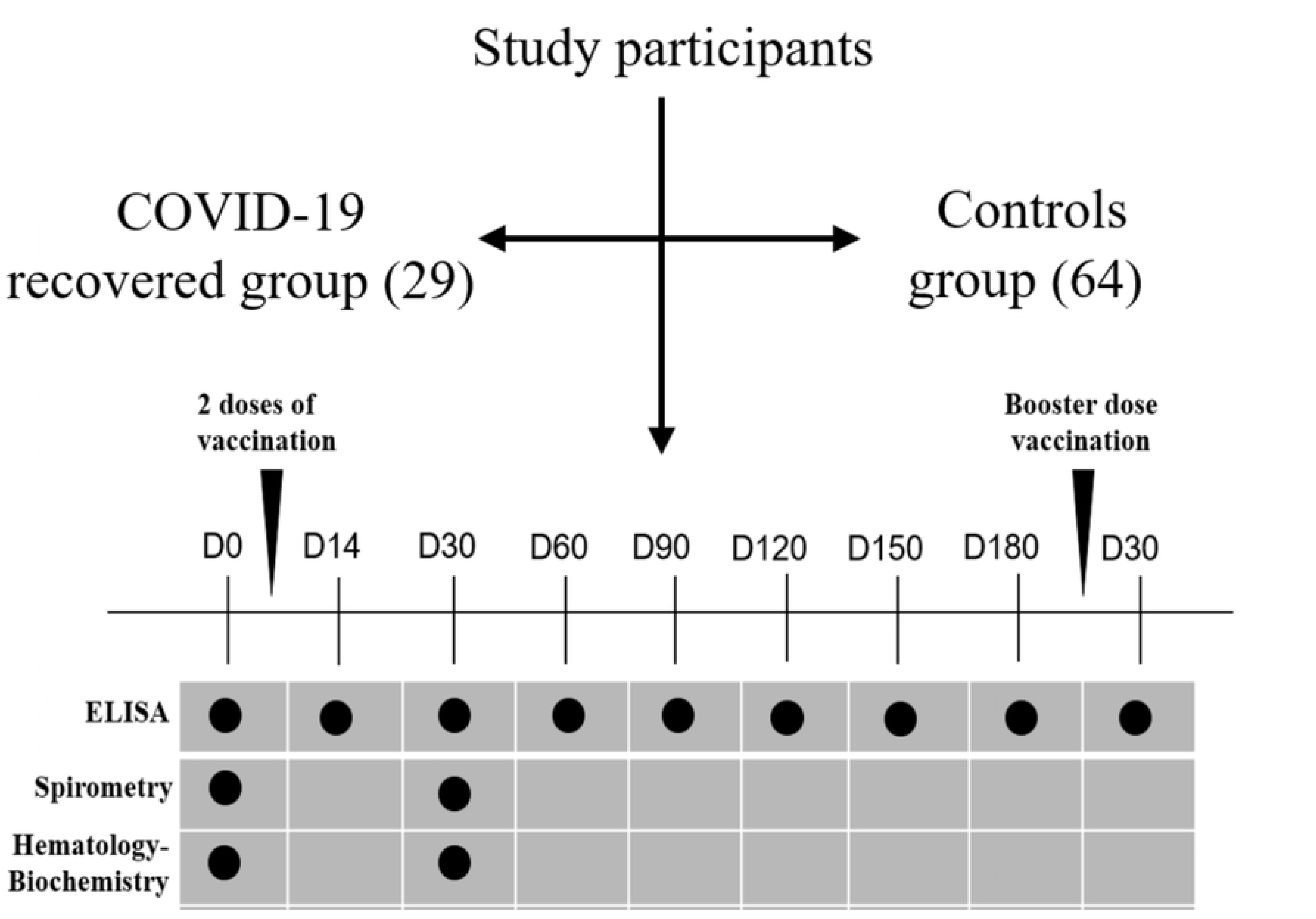
Flowchart of the study participants’ enrollment, follow-ups, and laboratory experiments conducted during the study period. ELISA, Spirometry and Biochemistry tests were performed at several day points throughout study period.

### 3.2 Sample Collection and laboratory tests

A total of 8 mL of blood was drawn, with 4 mL in a lavender-topped tube for biochemical and hematological tests, including complete blood count (CBC), random blood sugar (RBS), SGPT, and serum creatinine, and 4 mL in a red-topped tube for SARS-CoV-2 RBD-specific IgG quantification. The biochemical and hematological tests were performed twice: once before vaccination and again one month after the two vaccine doses.

### 3.3 Enzyme-Linked Immunosorbent Assay (ELISA)

Following the manufacturer’s instructions, we used the WANTAI SARS-CoV-2 IgG Elisa Kit (Quantitative) to analyze the IgG antibody response specific to RBD in sera collected from all of the participants measured at multiple time points **(Fig 1)**.

### 3.4 Pulmonary Functional Test (PFT)

A standard spirometry test was performed on each patient using a MicroQuark COSMED spirometer (The Metabolic Company, Italy) to measure their forced expiratory volume in the first second (FEV1), Forced Vital Capacity (FVC), FEV1/FVC ratio, Forced Expiratory Flow (FEF), Maximal Expiratory Flow (MEF) and Peak Expiratory Flow (PEF). The results were presented as a percentage of the predicted values comparing with the normal values for the Bangladeshi population. Values were chosen from the best of three attempts, and %predicted normal values were taken as >80% of normal (34).

Spirometry tests were first performed on the study participants. For those with pre- bronchodilation (BD) parameters below 80%, a bronchodilator (400 mcg salbutamol) was administered, and spirometry measurements were taken again afterward. The interval between pre- and post-BD spirometry measurements was 15 minutes. All PFTs were supervised by the same well-trained technician carried out at Ingenious PulmO-FIT Health Care Limited, Dhaka, Bangladesh. PFTs were supervised by the same well-trained technician carried out at Ingenious PulmO-FIT Health Care Limited, Dhaka, Bangladesh.

### 3.5 Statistical analysis

Continuous variables are reported as medians and interquartile ranges (IQR) and categorical variables are reported as frequencies and percentages. Demographics and clinical characteristics of the study participants were compared between groups using Kruskal-Wallis rank sum test and Fisher’s exact test as appropriate. The Wilcoxon signed-rank test was used to calculate p-values for the Pattern of PFT parameters %predicted value changes among bronchodilator recipients. Correlation analysis between SARS-CoV-2 IgG antibody titers and pulmonary function parameters was conducted using the Pearson correlation coefficient. The Pearson correlation test was also used to calculate p-values for comparing age specific distribution of SARS-CoV-2 IgG antibody titer and PFT parameters across the study population. Data processing, statistical analysis, and creation of figures were done with R statistical software (version 4.3.1). Regarding missing data, we planned to exclude participants for whom vaccine immune response status (different follow-up day points) could not be determined. For all tests, statistical significance was defined using a two-sided significance level of α=0.05 and p < 0.05 was considered statistically significant.

## 4. Results

### 4.1 Study population with demographic and clinical characteristics

The study enrolled a total of 93 participants: 29 who had experienced COVID-19 with varying severities (15 mild, 6 moderate, and 8 severe) and had recovered within the last 1-18 months, and 64 participants as controls. Participants in the study had a median age of 34 years (interquartile range: 18–67 years), and 67.7% of them were male. Among the recovered groups, the majority of the participants in moderate and

Data are presented as Median (IQR) and n (%). The Kruskal-Wallis rank sum test and Fisher’s exact test are used to calculate p values, respectively.

Severe groups (median age 47.5) were female. All the recovered groups had elevated BMIs; the moderately symptomatic group had the highest BMI (30 kg/m2) compared to the other mild, severe, and control groups. When comparing comorbidities, the most prevalent co-morbidities among the recovered group were diabetes, asthma, and hypertension. The most prevalent signs and symptoms among the study groups during enrollment were flu-like symptoms, muscle aches, and joint pain **(Table 1)**.

**Table 1.** Demographic and clinical characteristics of the study participants at the time of enrollment.

| Characteristics | Mild (n=15) | Moderate (n=6) | Severe (n=8) | Control (n=64) | P value |
| --- | --- | --- | --- | --- | --- |
| **Demography** |  |  |  |  |  |
| Age (years), median (IQR) | 33(18-67) | 47.5(34-65) | 47.5(23-65) | 32(19-64) | 0.267 |
| Male gender, number (%) | 11(73.3) | 1(16.6) | 3(37.5) | 48(75) | 0.011 |
| BMI (kg/m<sup>2</sup>), median (IQR) | 27(19-42) | 30(25-40) | 27(21-34) | 26(17-34) | 0.394 |
| Smoker, number (%) | 4(26.7) | 0(0.00) | 0(0.00) | 1(1.56) | 0.075 |
| **Physical examination (median, IQR)** |  |  |  |  |  |
| Heart rate (beats/min) | 82(62-105) | 100(80-118) | 98(85-120) | 87(63-120) | 0.283 |
| Respiratory rate | 22 (16-28) | 20 (18-24) | 23 (18-24) | 22 (14-93) | 0.442 |
| Systolic (mm/Hg) | 120 (100-140) | 125 (110-140) | 120 (110-160) | 120 (100-150) | 0.389 |
| Diastolic (mm/Hg) | 80 (80-90) | 80 (70-80) | 80 (80-90) | 80 (60-100) | 0.215 |
| **Co-morbidities (number)** |  |  |  |  |  |
| Hypertension | 2 | 2 | 5 | 7 | 0.004 |
| Chronic pulmonary disease | 4 | 2 | 1 | 3 | 0.066 |
| Asthma | 2 | 1 | 1 | 6 | 0.803 |
| Diabetes | 2 | 2 | 3 | 10 | 0.248 |
| **Clinical symptoms (number)** |  |  |  |  |  |
| Flu like symptoms | 5 | 3 | 4 | 25 | 0.813 |
| Chest pain | 2 | 0 | 2 | 1 | 0.024 |
| Muscle aches | 1 | 2 | 2 | 5 | 0.085 |
| Joint pain (arthralgia) | 1 | 3 | 3 | 7 | 0.017 |
| Fatigue/Malaise | 0 | 1 | 2 | 3 | 0.172 |
| Shortness of breathing | 4 | 1 | 0 | 3 | 0.038 |
| Conjunctivitis | 3 | 2 | 1 | 5 | 0.106 |
| Screen rash | 2 | 0 | 2 | 2 | 0.061 |
| **Family/ contact history (COVID-19)** | 10(83.3) | 5(83.3) | 7(87.5) | 6(9.3) | 0.000 |

Data are presented as Median (IQR) and n (%). The Kruskal-Wallis rank sum test and Fisher's exact test are used to calculate p values, respectively.

### 4.2 Hematological and biochemical parameters before and after vaccination

Prior to the vaccination and one month after receiving the two doses, collected blood samples were tested for hematology. Overall, no significant differences were observed between the study groups in biochemistry and hematology findings during pre- and post-vaccination periods. Hematology test results showed that none of the study group’s hemoglobin levels were abnormal. Moreover, no significant difference was observed in the overall white blood cell count (WBC) as well as differential count across the study groups. However, more elevated neutrophil counts were observed in the severe group than in the other recovered groups (severe 68% vs. mild 57% vs. moderate 56%). The moderate symptomatic group had the highest lymphocyte count (31%) compared to other groups. Other parameters, such as creatinine and RBS values, remained almost consistent across the study group **(Table 2)**.

**Table 2.** Biochemistry and hematology test results among the study groups before vaccination and after vaccination.

| Laboratory tests | Mild (n=15) |  | Moderate (n=6) |  | Severe (n=8) |  | Control (n=64) |  |
| --- | --- | --- | --- | --- | --- | --- | --- | --- |
|  | Pre-Vac | Post-Vac | Pre-Vac | Post-Vac | Pre-Vac | Post-Vac | Pre-Vac | Post-Vac |
| Hemoglobin (g/dL) | 12.9 (12.1- 14) | 13.3 (12.6- 14.4) | 12(11.2- 14.1) | 11.6 (11.3- 13) | 12.4 (11- 13.8) | 11.8(11- 13.3) | 14.3(12.1- 15.1) | 13.6(12.3- 14.6) |
| Total WBC (cmm) | 8000 (7250- 9000) | 9250 (8250- 10125) | 9500 (8000- 11000) | 7000 (7000- 7500) | 9250 (8375- 10750) | 9250 (8000- 10375) | 7500 (6500- 8625) | 7500 (6500- 8500) |
| Differential count (%) |  |  |  |  |  |  |  |  |
| Neutrophil | 57(50- 63) | 52(48-59) | 56(47-63) | 63 (53-66) | 68 (58-74) | 60 (54-64) | 58 (52-65) | 60 (54-64) |
| Lymphocyte count | 25 (3-36) | 33 (17-36) | 31 (26-37) | 27 (25-31) | 25 (21-27) | 32 (28-36) | 28 (25-35) | 31 (26-36) |
| Monocyte | 5 (3-7) | 5 (3-6) | 5 (2-7) | 4 (4-7) | 4 (3-6) | 4 (3-5) | 5 (2-8) | 5 (2-7) |
| Eosinophil | 3 (2-5) | 3 (2-6) | 3 (2-5) | 2 (1-3) | 2 (2-2) | 2 (1-5) | 4 (2-6) | 3 (2-5) |
| Total RBC (million/cmm) | 4.9(4.7-5.2) | 5(4.6- 5.1) | 4.2(3.9- 5) | 4.3(3.9- 4.4) | 4.9(4.3- 5.1) | 4.7(4.1- 5.1) | 5(4.7- 5.4) | 4.9(4.6- 5.2) |
| HCT/PCV% | 40.7 (38.4- 42.4) | 40.7 (39.2- 44.1) | 37.1 (35.4- 43.2) | 36.8 (36.1- 39.8) | 39.8 (35.5- 41.6) | 37.3 (35.5- 41.1) | 42.7 (37.6-45.6) | 41.0 (38.0-42.6) |
| MCV 1L | 83 (78-85) | 83.8 (77.6- 87.9) | 87 (85-91) | 87.6 (85.6- 90.2) | 81 (78-85) | 80.1 (77.4- 81.7) | 84 (79- 87) | 84.2 (79.2- 86.9) |
| MCH (pg) | 26.6 (24.6- 27.4) | 27.5 (24.9- 28.2) | 28.5 (27.8- 29.3) | 28.5 (26.3- 29.5) | 25.7 (24.8- 28.4) | 24.8(24.3- 27.3) | 27.9 (26.2- 29) | 27.8 (26- 29.5) |
| MCHC (g/dL) | 32.3 (31.6- 32.8) | 32.4 (32.2- 33) | 32.5 (31.5- 33) | 32.1 (30.7- 32.6) | 31.8 (31.4- 33.1) | 32.1 (30.9- 32.7) | 33.1 (32.3- 33.8) | 33.2 (32.3- 33.9) |
| RDW% | 14.1 (13.8- 15) | 14.1 (13.8- 14.7) | 14.3 (14.1- 16) | 14.2 (13.2- 14.3) | 14.5 (13.6- 16.2) | 14.3 (13.9- 15.3) | 13.6(13- 14.5) | 13.6 (13.1- 14.6) |
| Total platelet (count/cmm) X10<sup>4</sup> | 28(19-32) | 25(22.6-8.7) | 31.7(25.8-35) | 22(22-29) | 31(28.5-36.5) | 29(22.5- 33.5) | 28(23-32) | 26(22-30) |
| MPV 1L | 10.7 (10.2- 11.5) | 11.2 (10.4- 12) | 11 (10.7- 11.5) | 11.1 (10.2- 12.2) | 9.9 (9.7- 11.2) | 10.8 (10- 11.8) | 10.8 (10.2- 11.7) | 11.1 (10.6- 11.9) |
| PCT% | 0.3 (0.2- 0.3) | 0.3 (0.2- 0.3) | 0.3 (0.2- 0.3) | 0.2 (0.2- 0.2) | 0.2 (0.2- 0.3) | 0.3(0.2- 0.3) | 0.2 (0.2- 0.3) | 0.2 (0.2- 0.3) |
| PDW 1L | 13 (11.9- 14.7) | 14.2 (13.3- 15.9) | 14.5 (13.3- 15.1) | 12.9 (11.6- 15.9) | 11.8 (10.8- 14.1) | 13.2 (11.9- 15.6) | 13.1 (12.1- 16) | 13.9 (12.5- 15.8) |
| Biochemistry |  |  |  |  |  |  |  |  |
| RBS mmol/L | 5.1 (4.3- 5.5) | 5.6 (4.8- 6.2) | 6.2 (5.3- 6.9) | 7.2 (5.7- 7.5) | 7 (5.2- 9.2) | 6.9 (4.6- 10.7) | 5.2 (4.6- 5.9) | 5 (4.3- 6.5) |
| ALT (SGPT) U/L | 27 (23- 50) | 27 (23- 40) | 22 (19- 42) | 20 (18- 42) | 24 (18- 30) | 28 (22- 40) | 27 (19- 41) | 28 (22- 39) |
| Creatinine mg/dL | 0.7 (0.6- 0.8) | 0.7(0.6- 0.8) | 0.8 (0.6- 0.9) | 0.6 (0.5- 0.8) | 0.6(0.5- 0.7) | 0.7 (0.6- 0.8) | 0.8 (0.7- 0.9) | 0.8 (0.7- 0.9) |

Here, data are presented as median (IQR). "Pre-Vac" and "Post-Vac" represent the periods before and after vaccination, respectively.

### 4.3 Pulmonary function tests assessment

In all groups, the pulmonary functional test (PFT) was evaluated prior to vaccination and 30 days after two vaccination doses.

#### 4.3.1 Pulmonary function assessment before vaccination

All the participants were tested for PFT parameters for pulmonary functionality assessment. The study physician administered bronchodilators to 31 participants including 4 severe, 4 moderate, 8 mild, and 15 controls based on the FVC normal range by age, sex, and study physician discretion. The median %predicted score of FVC and FEV1 was below 80% in all recovered groups (mild to severe) before and after bronchodilation. However, median %predicted values of FEV1/FVC were found to be normal in all study groups **(Fig 2)**. Overall, recovered groups (moderate and severe) showed lower PFT %predicted scores compared to others during spirometry tests before vaccination **(Supplementary file S1)**.

**Fig. 2.**
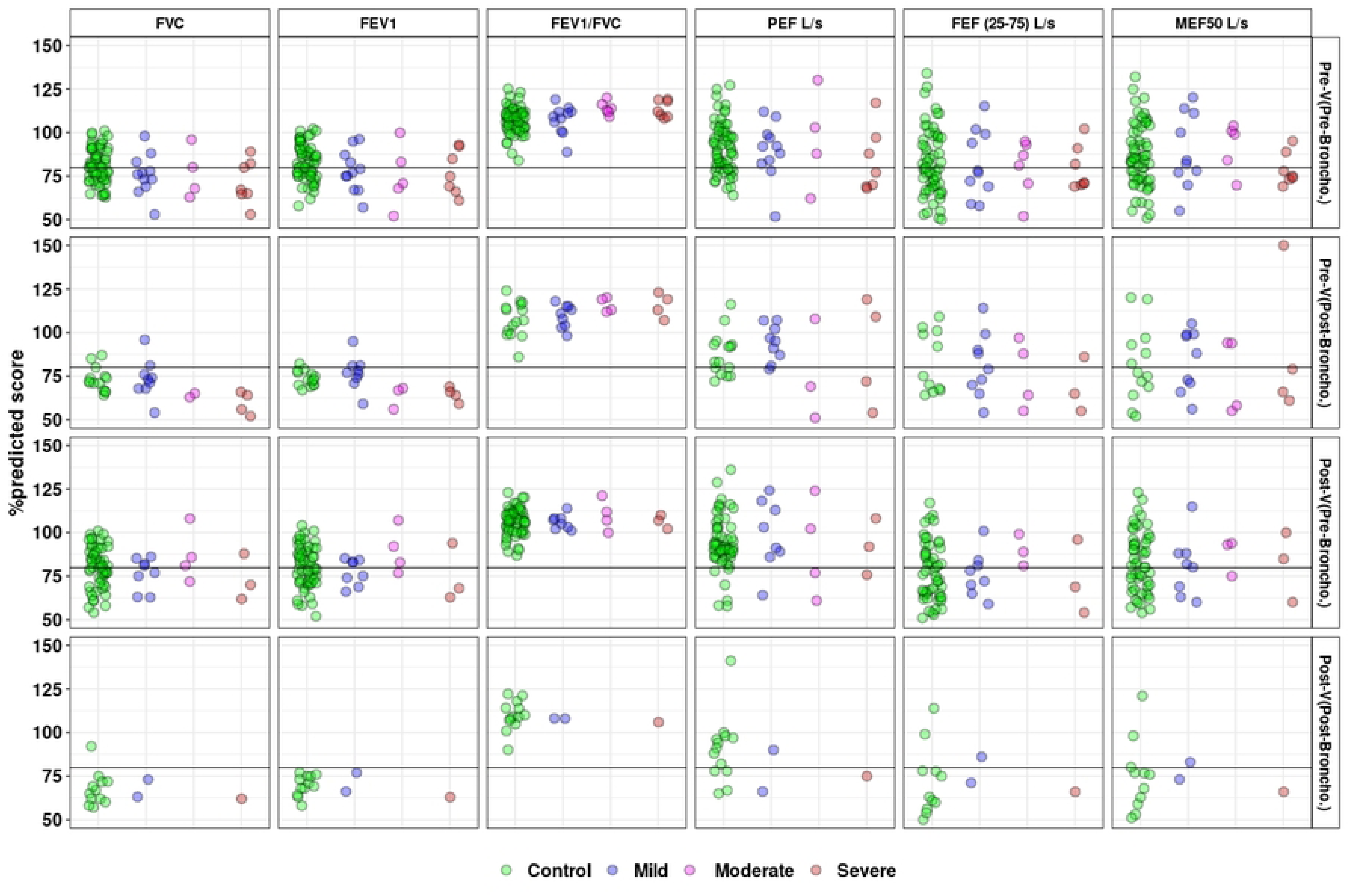
%predicted values for PFT parameters in the study groups were taken before and after vaccination. **These** parameters include Forced Vital Capacity (FVC), Forced Expiratory Volume in 1 Second (FEV1), FEV1/FVC ratio, Peak Expiratory Flow (PEF), Forced Expiratory Flow (FEF), and Maximal Expiratory Flow (MEF). Pre-V (Pre-Broncho) and Pre-V (Post-Broncho) represent PFT values obtained before and after the bronchodilation which was carried out before immunization. Post-V (Pre-Broncho) and Post-V (Post-Broncho) represent PFT values obtained before and after bronchodilation, which was performed after immunization.

Moreover, abnormal spirometry values were observed in several cases. Abnormal and below <80% FVC score was found in the 66% moderate group followed by 61.5% mild, 57.14% severe, and 23.4% control group. Almost similar patterns were observed for FEV1 values **(Fig 3)**. Additionally, we noted several paradoxical cases of bronchoconstriction in which PFT values declined even following bronchodilation. Of these, two mild, one moderate, and four severe—showed a decrease in the FVC %predicted score following bronchodilator **(Fig 3)**.

**Fig. 3.**
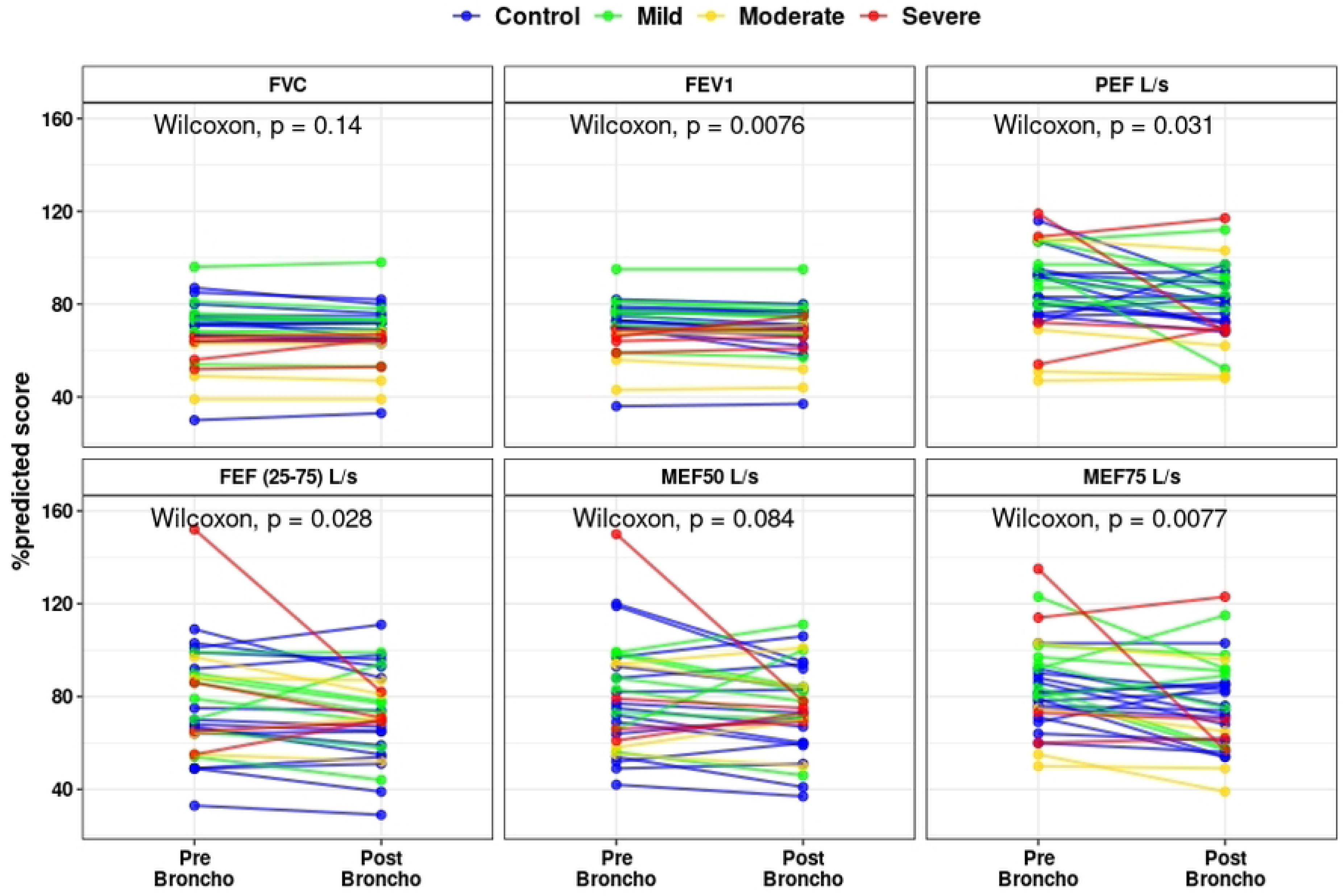
Pattern of PFT parameters %predicted value changes among the participants who received bronchodilator. Here, Pre-Broncho indicates the PFT %predicted values before the bronchodilator was given and Post-Broncho means PFT %predicted values after bronchodilator was administered. p-values were determined using the Wilcoxon signed-rank test.

#### 4.3.2 Pulmonary function assessment after vaccination

Additionally, we looked at the aforementioned PFT parameters 30 days after two doses of vaccination for 59 control, 10 mild, 4 moderate, and 3 severe participants. However, bronchodilation was given to a total of 15 participants, of whom 13 were control, 1 severe, and 1 mild based on the FVC normal range by age, sex, and study physician discretion. Pre-bronchodilation FVC mean %predicted scores found the lowest severe group in the severe group (73.3%) and mild (72.3%) group followed by the control group (80%) and moderate group (87%) **(Fig 2)**. In one case of mild and one case of severe participants, the FVC %predicted score even dropped **(Fig 3)**. The FEV1 %predicted score improved slightly but stayed below 80 among the 13 controls who were given bronchodilators. Likewise, in one mild case and one severe case, the FEV1 score did not rise above 80. Before bronchodilation, all study groups exhibited mean FEV1/FVC %predicted scores greater than 80. One moderate and one severe PEF L/s measurement subject did not show score improvement even after bronchodilation. Furthermore, even after bronchodilation, the predicted score for FEF (25–75)% for 10 control, 1 mild, and 1 severe participant remained below 80 **(Fig 3)**.

### 4.4 Duration and breadth of SARS-CoV-2 RBD specific antibody

This study evaluated the duration and breadth of SARS-CoV-2 RBD-specific antibody titer (IU/mL) before and after vaccination at different follow-up day points among the study groups. The seropositive cut-off value was ≥ 1 as per the kit manual.

Before vaccination, 47% of mild, 66.6% of moderate, and 100% of severe symptomatic individuals were seropositive where severe symptomatic group had the highest pre-vaccination antibody levels at 221.54 IU/mL followed by the mild, moderate, and control groups at 119.62 IU/mL, 51.74 IU/mL, and 24.94 IU/mL respectively. All study groups exhibited their highest antibody titers on day 14 following two vaccine doses. Among the recovered groups with varying symptom severity (mild to severe), antibody titers were significantly higher than the control group (p<0.005), where the severe group showed the highest titer (1418 IU/mL) **(Fig 4)**. However, a significant decline in antibody levels was observed between 90–120 days post-vaccination, particularly in the mild and control groups, though titers remained above baseline for up to 5–6 months within the study period. Interestingly, some symptomatic individuals showed increased titers at 3-4 months, suggesting potential breakthrough infections. Six months post-vaccination, a booster dose was administered. After 30 days post-booster vaccination, antibody titers significantly raised across all groups, with the severe group showing the highest increase (2368.01 IU/mL), about 1.5 times higher compared to the second dose **(Fig 4)**. Overall, antibody response findings suggest the importance of vaccination (booster dose) regardless of infection history.

**Fig. 4.**
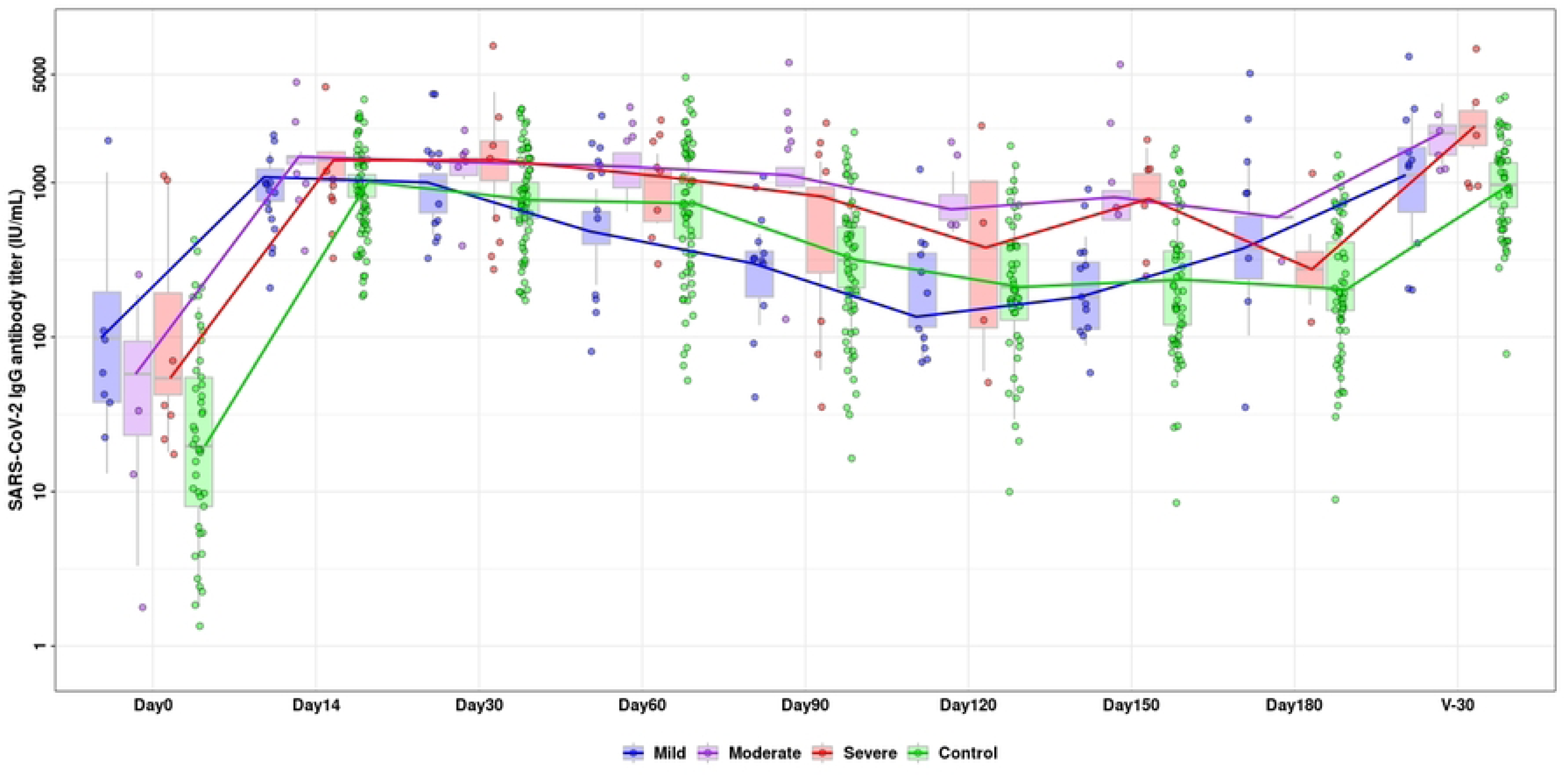
Duration and breadth of SARS-CoV-2 IgG antibody (IU/mL) response. Antibody titers measured over six months follow-ups following several day points including day14, day30, day60, day90, day120, day150, day 180 and a further 30 days after booster dose among the study groups.

### 4.5 Correlation between SARS-CoV-2 IgG antibody titer and Pulmonary Functional Parameter

This study examined the correlation between pulmonary function tests (PFTs) and SARS-CoV-2 IgG antibody titers. A negative correlation was observed between FVC and IgG antibody titers across all groups, with the severe group showing the strongest correlation (r =-0.575). Similarly, a negative correlation for FEV1 was found, with the severe group having the highest correlation (r =-0.454) **(Fig 5)**. FEV1/FVC showed a positive correlation across all groups, strongest in the mild to moderate group (r = 0.669, p = 0.003). While the control group showed weak negative correlations for PEF, FEF (25-75%), and MEF50%, the mild to moderate group displayed very weak positive correlations, and the severe group showed stronger positive correlations **(Fig 5)**.

**Fig. 5.**
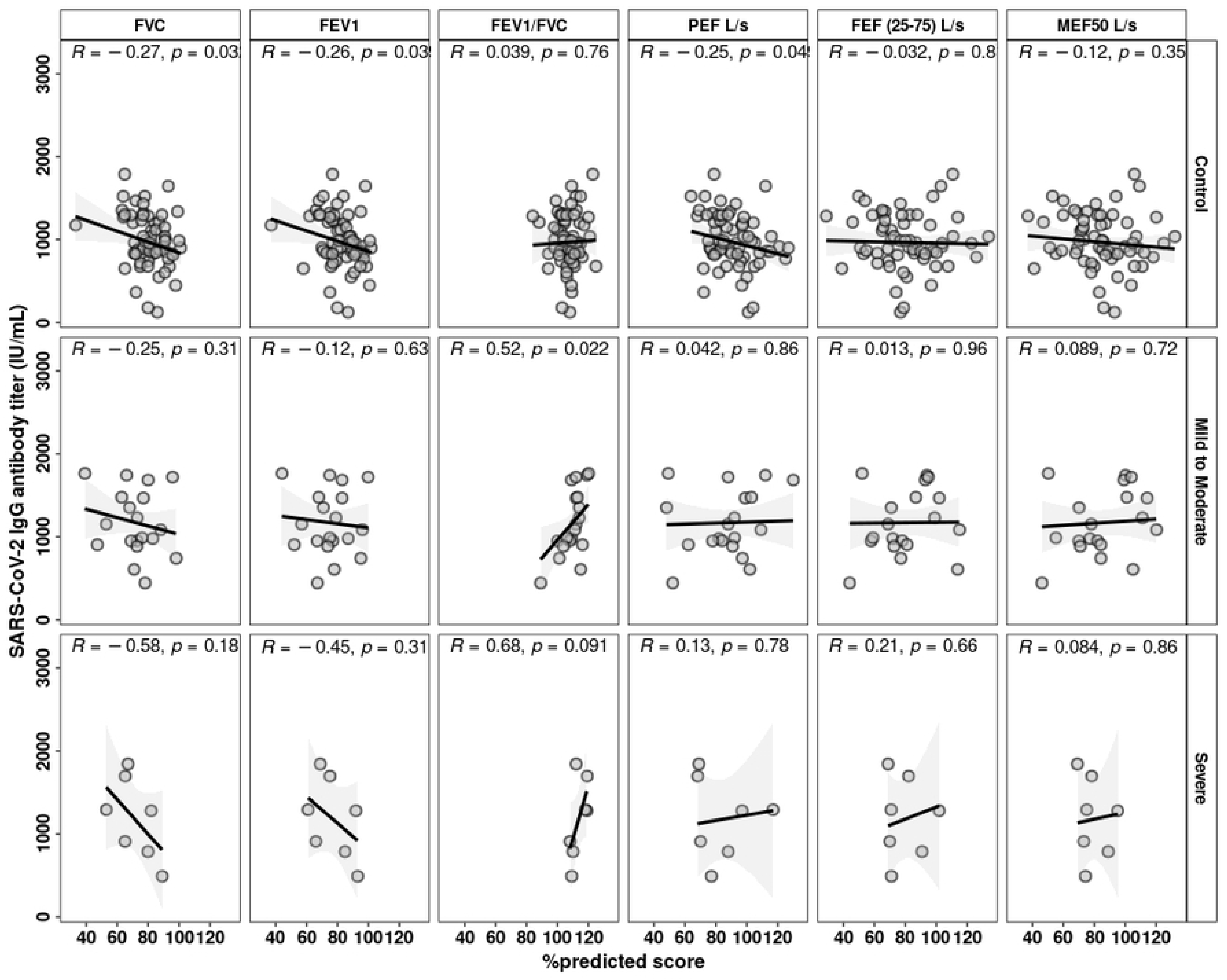
Correlation between pulmonary functional parameters (PFTs) and SARS-CoV-2 IgG antibody titers (IU/mL). Antibody titers were considered after 14 days of two doses of vaccination among the study groups including control, mild to moderate symptomatic group (MI to MO) and severe symptomatic group (SE) recovered from COVID-19. PFT parameters are presented in %predicted score value. The Pearson correlation method was used to calculate p-values.

We also found higher antibody titers among the individuals who showed abnormal PFT values compared to the individuals with normal PFT values **(Fig 6)**.

**Fig. 6.**
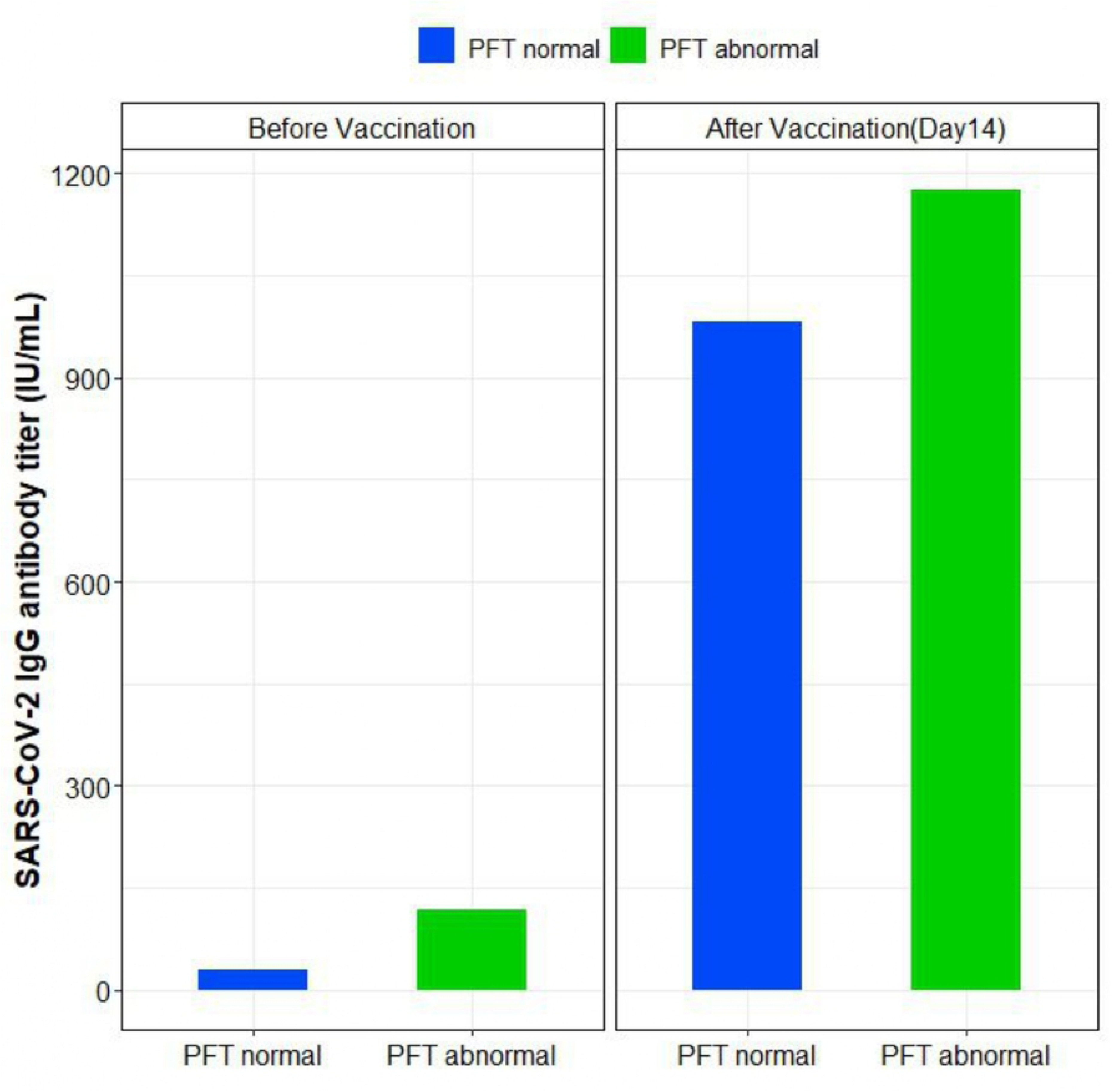
PFT parameters vs antibody titers. SARS-CoV-2 IgG antibody titer (IU/mL) among the study participants with PFT %predicted score with normal and abnormal values observed before and after vaccination.

### 4.6 Age specific distribution of SARS-CoV-2 IgG antibody titer and PFT parameters

All the participants were included in a subsequent analysis to look into correlations between IgG levels and age. A weak positive correlation was found between IgG levels and participant age at day 30 after the second vaccination dose, with a similar trend observed 30 days after the booster **(Fig 7)**.

**Fig. 7.**
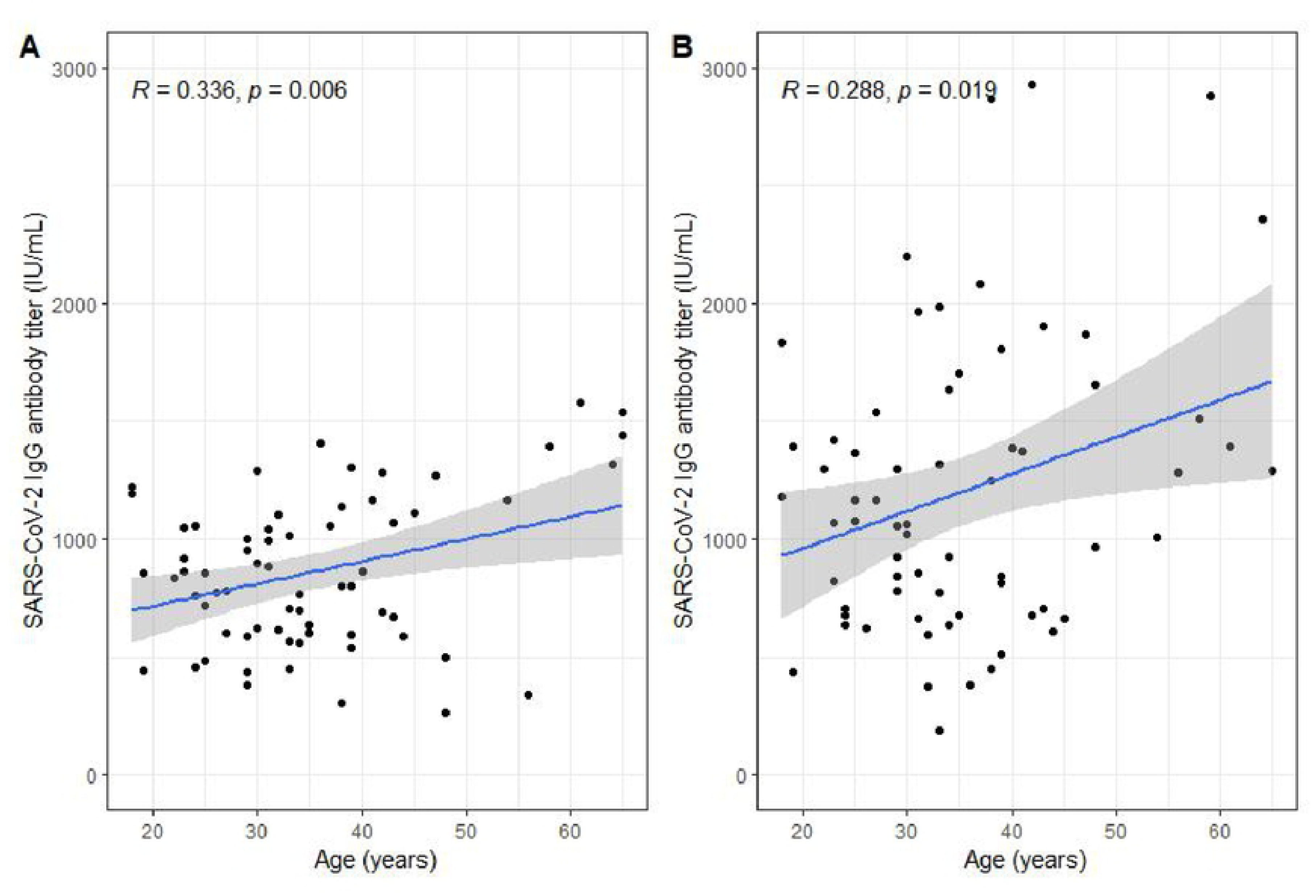
Age-specific correlation pattern of SARS-CoV-2 IgG antibody titer among the study groups. **A)** SARS-CoV-2 IgG antibody titer (IU/mL), 30 days after two doses of vaccination among the participants. **B)** SARS-CoV-2 IgG antibody titer (IU/mL), 30 days after booster dose (third dose) of vaccination among the study participants. The p-values were calculated using the Pearson correlation method.

Additionally, negative correlations were identified between age and PFT parameters except for FEF(25-75%) **(Fig 8)**.

**Fig. 8.**
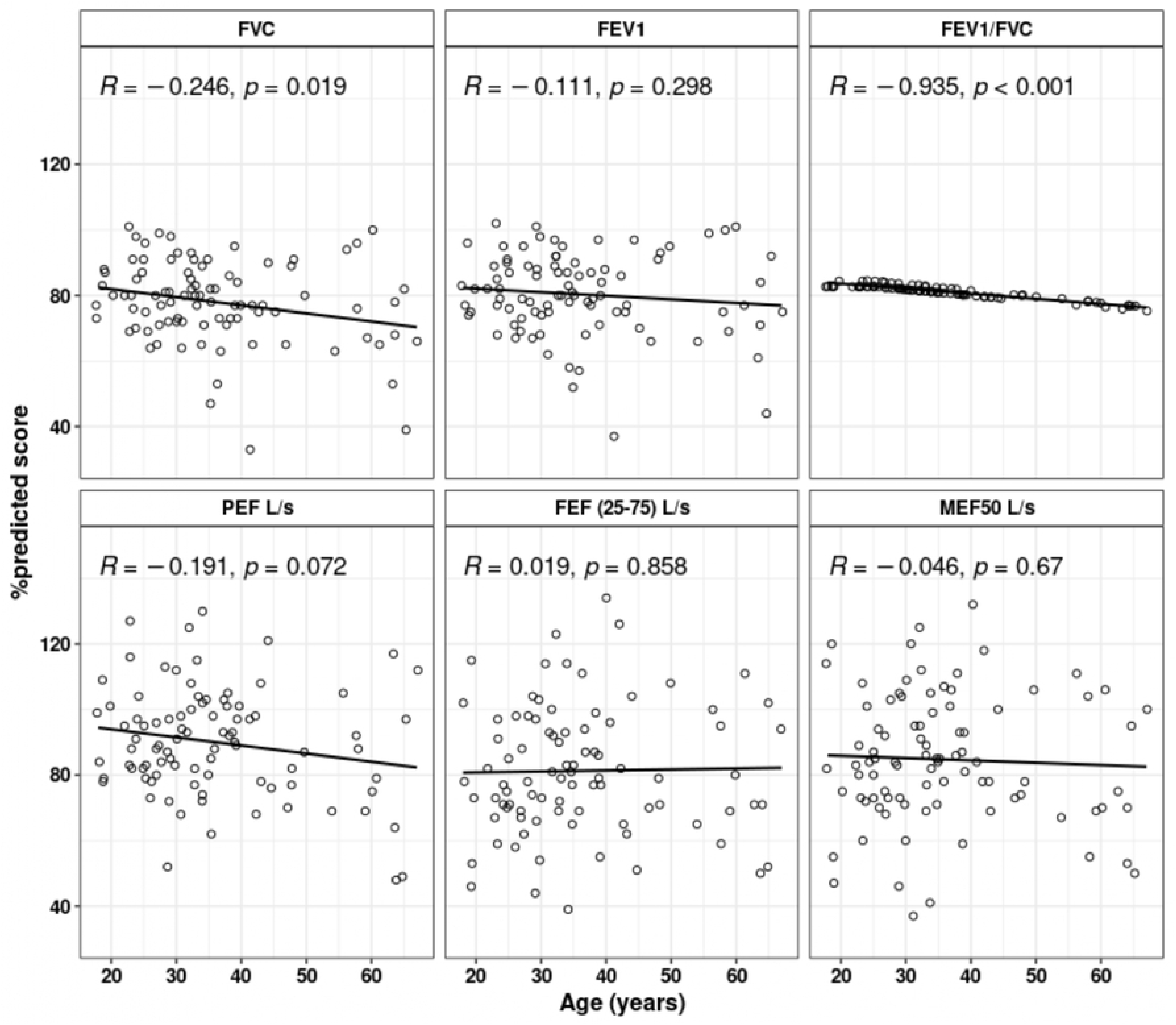
Correlation pattern between pulmonary functional test (PFT) parameters and age across the study population. Age in years and PFT parameters like FVC, FEV1, FEV1/FVC, PEF L/s, FEF (25–75) L/s and MEF50 L/s are shown in %predicted score value. p-values were determined using the Pearson correlation method.

## 5. Discussion

To the best of our knowledge, this is the first prospective study in Bangladesh assessing pulmonary function among individuals who have recovered from COVID-19 after experiencing mild to severe symptoms. Additionally, it includes a longitudinal evaluation of immune responses to investigate the duration and breadth of IgG antibody levels for up to six months following vaccination.

Importantly, this study highlights the important information about the long-term effects of COVID-19 on pulmonary function and the impact of vaccination, considering immune response (IgG antibody) among these aforementioned groups. Substantial geographic variation persists in the clinical presentations of symptomatic patients with SARS-CoV-2 infection, showcasing a diverse spectrum of symptoms and severity. The intricate interplay between COVID-19 infection history, varying disease severity, comorbidities’ impact on post-recovery pulmonary status, and vaccine-induced host immune responses poses a challenging yet vital avenue for research.

A nuanced understanding of COVID-19 recovery conditions, particularly in the context of pulmonary functional assessment, holds the potential to unveil therapeutic approaches and enhance the monitoring and evaluation of COVID-19 patients. The current study’s findings revealed that there were no significant differences in hematological and biochemical parameters observed before and after vaccination among the recovered group and the control group. However, neutrophil and lymphocyte counts were found to be higher among the moderate symptomatic group compared to other groups (Table 2). In addition to having higher neutrophil and platelet counts overall, the COVID-19 infection history with moderate disease severity group also had a higher MCV and MCH differential count, which was consistent with previous study findings (35,36).

While observing the pulmonary function assessment, the symptomatic group had the lowest mean %predicted score for FVC and FEV1. When we analyzed the mean FVC% projected scores, the moderate group experienced the lowest score (65.5%), followed by the severe group (71%). Moreover, FVC% predicted scores of below 80% were found in 61.5% of the mild, 66.6% of the moderate, and 57.14% of the severe group. However, compared to previous studies where pulmonary function was observed three to six months after symptom onset, the current study found a higher number of abnormal pulmonary function cases (37,38). When bronchodilators were given, FVC %predicted scores improved in 1 of 4 moderate, 4 of 8 mild, and 11 of 15 control group participants, but none in the severe group. Surprisingly, FVC scores declined in all severe cases and in some mild (2), moderate (2), and control (3) participants after bronchodilation **(Fig 3)**. Such paradoxical bronchoconstriction with respiratory distress was reported in several studies (39,40). Bronchodilators are prescribed to relax airway smooth muscle and improve airflow. Paradoxical bronchoconstriction with resulting decreased airflow occurs in some patients even after administration of bronchodilators. Paradoxical bronchoconstriction with resulting decreased airflow occurs in some patients after administration of bronchodilators.

Allergic reaction to inert constituents such as emulsifying agents, propellants, preservatives or even contaminants, inhalational technique or the characteristics of the solution (e.g., pH), β-receptor polymorphisms can contribute to paradoxical response (41–43). However, the exact mechanism is still not clear where this paradoxical acute bronchoconstriction remains to be determined.

Similar findings were observed for FEV1 %predicted scores. However, all groups showed %predicted score above 80 for FEV1/FVC. Additionally, PEF L/s and FEF (25-75%) showed no discernible differences. Moreover, %predicted scores for PEF and FEF (25-75%), were found higher among the symptomatic group, though the results were higher than the previous study findings (44). Such variations may be due to geographic and demographic differences, and the inconsistent definition of normality for pulmonary function parameters may also contribute to such findings. Due to the presence of prior comorbidities associated with COVID-19, defining pulmonary sequela is also quite challenging.

Overall, we observed lower FVC, FEV1, and PEF %predicted values with a disease severity history ranging from mild to severe, which corroborated the previous study findings compared to the control group (38,44–46). This suggests that following recovery from COVID-19, the majority of patients with a symptomatic history have pulmonary functional abnormalities that have been present for at least 6 to 12 months. However, some studies have found little to no differences in pulmonary function as determined by PFT parameters before and after COVID-19 [12], which may be explained by the study population’s diversity and the inclusion of hospitalized patients (47,48).

Additionally, the higher BMI of patients with symptoms may be one of the reasons to explain this. Moreover, in this study, recovered patients who experienced mild to moderate symptoms commonly reported cough, arthralgia, fatigue/malaise, shortness of breath, and anaphylactic reactions. Similar symptoms were also noted in other studies (49,50).

We observed a comparatively negative correlation of FVC and FEV1 with antibody titers among symptomatic groups. Similar findings were observed in previous studies where these parameters were significantly reduced among the group with previous severe symptomatic infection history (51).

After respiratory viral infections, such as COVID-19, lung damage and chronic pathology are mostly caused by prolonged and/or dysregulated immune responses, especially involving CD8+ T cells. When CD69+CD103− T cells are apparent in elderly COVID-19 convalescents, they release more cytotoxic and inflammatory compounds, commonly which is associated with deteriorating pathology and reduced lung function (52). Reduced lung function and delayed pulmonary recovery have also been linked to elevated SARS-CoV-2 S-specific IgG antibody titers and elevated levels of inflammatory markers, such as IL-6 (53). Interestingly, higher levels of IgG following vaccination, which reflect robust activation of the humoral immune system, are driven by memory B cells primed during initial exposure or vaccination. These cells ensure long-term immunity by rapidly producing antibodies upon re-exposure to the pathogen. However, excessive or dysregulated antibody responses can exacerbate inflammatory pathways or interact with other immune cells (such T cells), which could potentially contribute to post-infection respiratory issues, especially in circumstances of prior severe infection (54).

rIt is interesting to note that we observed correlations between age group and PFT values where FVC and FEV1 negatively correlated with the increased age group. Previous study also reported such correlation with increased age group (55). There is a positive correlation between age and increased antibody levels, as shown by the pattern of increasing antibody titers with age. In our study, the elderly age group demonstrated higher antibody titers after vaccination, which is comparable to a prior study in which the elderly group’s antibody titers were found to be higher following a naturally occurring COVID-19 infection (56). However, several studies have revealed that young people exhibit a higher antibody response to vaccination than elderly people (57). According to our study, this circumstance may be connected to a history of infections that caused disease severity, mostly in the elderly population, and the study population in this study was ≥ 18 years old.

Vaccination is without a doubt the best strategy to combat COVID-19, where effective vaccines, correct doses, and safe administration are essential. While cellular immunity plays a crucial role in virus clearance, humoral immunity also offers protection against viral infections, making it one of the fundamental targets of vaccination strategy (24). Studies suggest that elevated SARS-CoV-2 antibody levels may be linked to disease severity, potentially due to antibody-dependent enhancement, a phenomenon observed in dengue virus infections (58–60). A recently published work suggested vaccination against SARS-CoV-2 appears to be associated with the protection against subclinical infection. This study also reported that healthy subjects may exhibit respiratory distress who had an infection where vaccination might play a significant protective role, urging the importance of universal vaccination (61). Additionally, it has been reported that patients with more severe COVID-19 had stronger antibody responses. Nevertheless, the duration and breadth of the antibody response may differ in distinct populations, and it is still unclear how long the COVID-19-recovered individuals’ antibody response will persist against vaccination. In this study, the serological analysis was conducted on average six months after the two doses of vaccination. This result aligns with earlier findings that antibody titer was found to be higher among the recovered group, and the antibody level persisted for several months following two doses of vaccination (62,63). In addition, we noticed that the IgG level decline over time was more rapid in people who had never experienced COVID-19 with symptoms, corroborated with previous studies (64–66).

After two doses of vaccination, previously infected people with varying disease severity (from mild to severe) displayed a noticeable increase in antibody titers and were able to maintain comparatively high IgG antibody titer levels for a while longer than the control population, which is similar to previous study findings (67).

The importance of vaccination is emphasized by the finding that IgG antibody titers in all groups were higher after vaccination than their pre-vaccination baseline antibody titers, even after a six-month follow-up. Similar results from a 10-month follow-up study where the antibody titer was found to be higher than before vaccination seropositivity (68). Our study, however, showed that antibody titer levels in all groups dropped steadily within 3–4 months after two doses of vaccination, regardless of the infection history, indicating that further vaccinations might be advantageous even for those who had the infection before. These findings suggest the importance of vaccination dose as the antibody titer sharply declined after a certain period among all groups. This study has unequivocally demonstrated that the administration of a booster dose is highly effective in inducing a robust increase in antibody response within a short period. This finding also underlines the critical importance of vaccination against emerging strains for everyone, regardless of one’s previous infection history to protect themselves and their community against the spread and ongoing threat of this virus.

This study has several limitations. Some participants were lost to follow-up due to relocation or pregnancy. We did not measure Total Lung Capacity (TLC) or the lung’s diffusing capacity for carbon monoxide (DLCO) while performing functional assessment. Additionally, breakthrough infections were not tested, and no pseudo-neutralization assay was performed to assess the functional antibody response.

However, our study provides some crucial evidence to date among the Bangladeshi population on pulmonary functional activity upon recovery from COVID-19 as well as their antibody response following vaccination, which urges the importance of vaccination regardless of infection history and the importance of lung function assessment following recovery. Further investigation is warranted to delineate the temporal onset of persistent pulmonary symptoms and assess pulmonary functionality in individuals recovering from COVID-19 disease severities. The enduring pulmonary sequelae observed underscore the imperative for extended COVID-19 treatment strategies, emphasizing the prevention of long-term complications. Additionally, evaluating the immune response post-vaccination in COVID-19-recovered individuals becomes crucial to determining the necessity for vaccine doses and vaccination against emerging strains.

Although other studies have demonstrated a relationship between the severity of COVID-19 infection and high SARS-CoV-2 antibody titers, it appears that pulmonary functional abnormalities may exist in people who have previously experienced a moderate to severe symptomatic COVID-19 infection, even up to one year after recovery. Also, regardless of a person’s history with COVID-19, multiple vaccination doses might be advised. Overall, knowing how host immune responses relate to the severity of the illness and the persistence of pulmonary functional impairment in COVID-19 patients will help develop novel therapeutics and immunization strategies in the future pan-demic, such as the dosage and timing of the vaccination. Further research is therefore required to fully understand the potential antibody response with neutralization ability following vaccination against emerging strains as well as to monitor pulmonary function with a focus on long-term COVID risk, especially for the elderly population to ensure healthy respiratory health.

## 6. Conclusion

In conclusion, our findings highlight the significance of evaluating pulmonary function in individuals who recovered from COVID-19 and those who may have asymptomatic infections, to maintain long-term respiratory health. Elevated IgG antibody titers were observed following two doses of vaccination, particularly in those recovered persons with a history of severe and moderate infections, however, these titers decreased after a few months across all the study groups, emphasizing the importance of a further vaccination (booster doses) regardless of infection history. Overall, these findings highlight the need for comprehensive post-recovery care and immunization schemes for immune protection against emerging strains in promoting long-term respiratory health and effective pandemic preparation.

## Data Availability

All data produced in the present study are available upon reasonable request to the authors

## 7. Acknowledgements

We would like to thank officials and healthcare workers at Ingenious Health Care Limited, Dhaka, Bangladesh, and Anwar Khan Medical College, Dhaka, Bangladesh. Also, we would like to thank the National Institute of Health including the Fogarty International Center, Training Grant in Vaccine Development, and Public Health ((TW005572) for providing support to ABS, MTH and others.

## 9 Supporting information

**File S1 %Predicted scores of Pulmonary Function Test (PFT) parameters among the study participants**. PFT parameters including forced vital capacity (FVC), forced expiratory volume in 1 second (FEV1), FEV1/FVC, peak expiratory flow (PEF), The maximal expiratory flow (MEF), forced expiratory flow (FEF) with pre and post bronchodilation %predicted scores both before and after vaccination among the mild, moderate, severe and control study groups. %predicted score shown in median value with interquartile range (IQR).

## 8. References

1. Zhu N, Zhang D, Wang W, Li X, Yang B, Song J, et al. A Novel Coronavirus from Patients with Pneumonia in China, 2019. N Engl J Med. 2020 Feb 20;382(8):727–33.

2. Valencia DN. Brief Review on COVID-19: The 2020 Pandemic Caused by SARS-CoV-2. Cureus. 2020 Mar 24;12(3):e7386.

3. Tackling the COVID-19 pandemic: The Bangladesh perspective - PMC [Internet]. [cited 2024 Oct 3]. Available from: https://www.ncbi.nlm.nih.gov/pmc/articles/PMC7582102/

4. Hernandez Acosta RA, Esquer Garrigos Z, Marcelin JR, Vijayvargiya P. COVID-19 Pathogenesis and Clinical Manifestations. Infect Dis Clin North Am. 2022 Jun;36(2):231–49.

5. Yao XH, Li TY, He ZC, Ping YF, Liu HW, Yu SC, et al. [A pathological report of three COVID-19 cases by minimal invasive autopsies]. Zhonghua Bing Li Xue Za Zhi. 2020 May 8;49(5):411–7.

6. Bradley BT, Maioli H, Johnston R, Chaudhry I, Fink SL, Xu H, et al. Histopathology and ultrastructural findings of fatal COVID-19 infections in Washington State: a case series. Lancet. 2020 Aug 1;396(10247):320–32.

7. Liu K, Zhang W, Yang Y, Zhang J, Li Y, Chen Y. Respiratory rehabilitation in elderly patients with COVID-19: A randomized controlled study. Complement Ther Clin Pract. 2020 May;39:101166.

8. Hui DS, Joynt GM, Wong KT, Gomersall CD, Li TS, Antonio G, et al. Impact of severe acute respiratory syndrome (SARS) on pulmonary function, functional capacity and quality of life in a cohort of survivors. Thorax. 2005 May;60(5):401–9.

9. Xie L, Liu Y, Xiao Y, Tian Q, Fan B, Zhao H, et al. Follow-up study on pulmonary function and lung radiographic changes in rehabilitating severe acute respiratory syndrome patients after discharge. Chest. 2005 Jun;127(6):2119–24.

10. Lindahl ALL, Aro M, Reijula J, Puolanne M, Mäkelä MJ, Vasankari T. Persisting symptoms common but inability to work rare: a one-year follow-up study of Finnish hospitalised COVID-19 patients. Infectious Diseases. 2023;55(12):821–30.

11. Salem AM, Al Khathlan N, Alharbi AF, Alghamdi T, AlDuilej S, Alghamdi M, et al. The Long-Term Impact of COVID-19 Pneumonia on the Pulmonary Function of Survivors. Int J Gen Med. 2021;14:3271–80.

12. Liang L, Yang B, Jiang N, Fu W, He X, Zhou Y, et al. Three-month Follow-up Study of Survivors of Coronavirus Disease 2019 after Discharge. J Korean Med Sci. 2020 Nov 25;35(47):e418.

13. Nishiga M, Wang DW, Han Y, Lewis DB, Wu JC. COVID-19 and cardiovascular disease: from basic mechanisms to clinical perspectives. Nat Rev Cardiol. 2020 Sep;17(9):543–58.

14. Tay MZ, Poh CM, Rénia L, MacAry PA, Ng LFP. The trinity of COVID-19: immunity, inflammation and intervention. Nat Rev Immunol. 2020 Jun;20(6):363–74.

15. Merad M, Martin JC. Pathological inflammation in patients with COVID-19: a key role for monocytes and macrophages. Nat Rev Immunol. 2020 Jun;20(6):355–62.

16. Sariol A, Perlman S. Lessons for COVID-19 Immunity from Other Coronavirus Infections. Immunity. 2020 Aug 18;53(2):248–63.

17. Braun J, Loyal L, Frentsch M, Wendisch D, Georg P, Kurth F, et al. SARS-CoV-2-reactive T cells in healthy donors and patients with COVID-19. Nature. 2020 Nov;587(7833):270–4.

18. Irsara C, Egger AE, Prokop W, Nairz M, Loacker L, Sahanic S, et al. Clinical validation of the Siemens quantitative SARS-CoV-2 spike IgG assay (sCOVG) reveals improved sensitivity and a good correlation with virus neutralization titers. Clinical Chemistry and Laboratory Medicine (CCLM). 2021 Jul 1;59(8):1453–62.

19. Vashist SK. In Vitro Diagnostic Assays for COVID-19: Recent Advances and Emerging Trends. Diagnostics (Basel). 2020 Apr 5;10(4):202.

20. Batra M, Tian R, Zhang C, Clarence E, Sacher CS, Miranda JN, et al. Role of IgG against N-protein of SARS-CoV2 in COVID19 clinical outcomes. Sci Rep. 2021 Feb 10;11(1):3455.

21. Ripperger TJ, Uhrlaub JL, Watanabe M, Wong R, Castaneda Y, Pizzato HA, et al. Orthogonal SARS-CoV-2 Serological Assays Enable Surveillance of Low-Prevalence Communities and Reveal Durable Humoral Immunity. Immunity. 2020 Nov 17;53(5):925–933.e4.

22. Yao L, Wang GL, Shen Y, Wang ZY, Zhan BD, Duan LJ, et al. Persistence of Antibody and Cellular Immune Responses in Coronavirus Disease 2019 Patients Over Nine Months After Infection. The Journal of Infectious Diseases. 2021 Aug 15;224(4):586–94.

23. Figueiredo-Campos P, Blankenhaus B, Mota C, Gomes A, Serrano M, Ariotti S, et al. Seroprevalence of anti-SARS-CoV-2 antibodies in COVID-19 patients and healthy volunteers up to 6 months post disease onset. Eur J Immunol. 2020 Dec;50(12):2025–40.

24. Liu L, Wei Q, Lin Q, Fang J, Wang H, Kwok H, et al. Anti-spike IgG causes severe acute lung injury by skewing macrophage responses during acute SARS-CoV infection. JCI Insight. 2019 Feb 21;4(4):e123158, 123158.

25. Tenforde MW, Patel MM, Ginde AA, Douin DJ, Talbot HK, Casey JD, et al. Effectiveness of SARS-CoV-2 mRNA Vaccines for Preventing Covid-19 Hospitalizations in the United States. medRxiv. 2021 Jul 8;2021.07.08.21259776.

26. Lo Sasso B, Agnello L, Giglio RV, Gambino CM, Ciaccio AM, Vidali M, et al. Longitudinal analysis of anti-SARS-CoV-2 S-RBD IgG antibodies before and after the third dose of the BNT162b2 vaccine. Sci Rep. 2022 May 23;12(1):8679.

27. Keshavarz B, Richards NE, Workman LJ, Patel J, Muehling LM, Canderan G, et al. Trajectory of IgG to SARS-CoV-2 After Vaccination With BNT162b2 or mRNA-1273 in an Employee Cohort and Comparison With Natural Infection. Front Immunol. 2022;13:850987.

28. Altawalah H. Antibody Responses to Natural SARS-CoV-2 Infection or after COVID-19 Vaccination. Vaccines (Basel). 2021 Aug 16;9(8):910.

29. Wu J, Liang B, Chen C, Wang H, Fang Y, Shen S, et al. SARS-CoV-2 infection induces sustained humoral immune responses in convalescent patients following symptomatic COVID-19. Nat Commun. 2021 Mar 22;12(1):1813.

30. Garcia-Beltran WF, Lam EC, Astudillo MG, Yang D, Miller TE, Feldman J, et al. COVID-19-neutralizing antibodies predict disease severity and survival. Cell. 2021 Jan 21;184(2):476–488.e11.

31. (PDF) SARS-CoV-2 Antibody Responses Are Correlated to Disease Severity in COVID-19 Convalescent Individuals [Internet]. [cited 2024 Oct 3]. Available from: https://www.researchgate.net/publication/347045892_SARS-CoV-2_Antibody_Responses_Are_Correlated_to_Disease_Severity_in_COVID-19_Convalescent_Individuals

32. ResearchGate [Internet]. [cited 2025 Jan 13]. (PDF) Covid-19 on the Economy & Society impact of Bangladesh. Available from: https://www.researchgate.net/publication/358265227_Covid-19_on_the_Economy_Society_impact_of_Bangladesh

33. COVID-19 clinical management: living guidance, 25 January 2021 [Internet]. [cited 2024 Oct 3]. Available from: https://iris.who.int/handle/10665/338882

34. Stanojevic S, Wade A, Stocks J. Reference values for lung function: past, present and future. Eur Respir J. 2010 Jul;36(1):12–9.

35. Zhang L, Huang B, Xia H, Fan H, Zhu M, Zhu L, et al. Retrospective analysis of clinical features in 134 coronavirus disease 2019 cases. Epidemiol Infect. 148:e199.

36. Soraya GV, Ulhaq ZS. Crucial laboratory parameters in COVID-19 diagnosis and prognosis: An updated meta-analysis. Med Clin (Engl Ed). 2020 Aug 28;155(4):143–51.

37. Liao X, Wang Y, He Z, Yun Y, Hu M, Ma Z, et al. Three-Month Pulmonary Function and Radiological Outcomes in COVID-19 Survivors: A Longitudinal Patient Cohort Study. Open Forum Infect Dis. 2020 Nov 14;8(9):ofaa540.

38. Polese J, Ramos AD, Moulaz IR, Sant’Ana L, Lacerda BS de P, Soares CES, et al. Pulmonary function and exercise capacity six months after hospital discharge of patients with severe COVID-19. Braz J Infect Dis. 2023;27(4):102789.

39. Magee JS, Pittman LM, Jette-Kelly LA. Paradoxical Bronchoconstriction with Short-Acting Beta Agonist. Am J Case Rep. 2018 Oct 9;19:1204–7.

40. Schissler AJ, Celli BR. Prevalence of paradoxical bronchoconstriction after inhaled albuterol. Respir Med. 2018 Aug;141:100–2.

41. (PDF) Incidence of acute decreases in peak expiratory flow following the use of metered-dose inhalers in asthmatic patients. ResearchGate [Internet]. 2024 Oct 22 [cited 2024 Dec 31]; Available from: https://www.researchgate.net/publication/15486382_Incidence_of_acute_decreases_in_peak_expiratory_flow_following_the_use_of_metered-dose_inhalers_in_asthmatic_patients

42. Wilkinson JR, Roberts JA, Bradding P, Holgate ST, Howarth PH. Paradoxical bronchoconstriction in asthmatic patients after salmeterol by metered dose inhaler. BMJ. 1992 Oct 17;305(6859):931–2.

43. Paradoxical bronchospasm associated with the use of inhaled beta agonists [Internet]. [cited 2024 Dec 31]. Available from: https://www.periodicos.capes.gov.br/index.php/acervo/buscador.html?task=detalhes&id=W2144298901

44. Huang Y, Tan C, Wu J, Chen M, Wang Z, Luo L, et al. Impact of coronavirus disease 2019 on pulmonary function in early convalescence phase. Respir Res. 2020 Jun 29;21(1):163.

45. Riou M, MarcoT C, Oulehri W, Enache I, Pistea C, Chatron E, et al. Respiratory follow-up after hospitalization for COVID-19: Who and when? Eur J Clin Invest. 2021 Aug;51(8):e13603.

46. Qin W, Chen S, Zhang Y, Dong F, Zhang Z, Hu B, et al. Diffusion capacity abnormalities for carbon monoxide in patients with COVID-19 at 3-month follow-up. Eur Respir J. 2021 Jul;58(1):2003677.

47. Lewis KL, Helgeson SA, Tatari MM, Mallea JM, Baig HZ, Patel NM. COVID-19 and the effects on pulmonary function following infection: A retrospective analysis. EClinicalMedicine. 2021 Sep;39:101079.

48. Cherrez-Ojeda I, Sanchez-Angarita E, Vanegas E, Farfán Bajaña MJ, Robles-Velasco K, Osorio MF, et al. Pulmonary Evaluation of Post-COVID-19 Patients: An Ecuadorian Experience. J Community Hosp Intern Med Perspect. 2022;12(2):30–4.

49. Carfì A, Bernabei R, Landi F, Gemelli Against COVID-19 Post-Acute Care Study Group. Persistent Symptoms in Patients After Acute COVID-19. JAMA. 2020 Aug 11;324(6):603–5.

50. Kanne JP, Little BP, Schulte JJ, Haramati A, Haramati LB. Long-term Lung Abnormalities Associated with COVID-19 Pneumonia. Radiology. 2023 Feb;306(2):e221806.

51. Guler SA, Ebner L, Aubry-Beigelman C, Bridevaux PO, Brutsche M, Clarenbach C, et al. Pulmonary function and radiological features 4 months after COVID-19: first results from the national prospective observational Swiss COVID-19 lung study. Eur Respir J. 2021 Apr;57(4):2003690.

52. Goplen NP, Cheon IS, Sun J. Age-Related Dynamics of Lung-Resident Memory CD8+ T Cells in the Age of COVID-19. Front Immunol [Internet]. 2021 Mar 29 [cited 2024 Dec 31];12. Available from: https://www.frontiersin.org/journals/immunology/articles/10.3389/fimmu.2021.636118/full

53. Wang Z, Wang S, Goplen NP, Li C, Cheon IS, Dai Q, et al. PD-1hi CD8+ resident memory T cells balance immunity and fibrotic sequelae. Sci Immunol. 2019 Jun 14;4(36):eaaw1217.

54. Proinflammatory IgG Fc structures in patients with severe COVID-19 | Nature Immunology [Internet]. [cited 2024 Dec 31]. Available from: https://www.nature.com/articles/s41590-020-00828-7

55. Afsin E, Demirkol ME. Post-COVID Pulmonary Function Test Evaluation. Turk Thorac J. 2022 Nov 1;23(6):387–94.

56. Petrovic V, Vukovic V, Patic A, Marković M, Ristić M. Immunogenicity of BNT162b2, BBIBP-CorV and Gam-COVID-Vac vaccines and immunity after natural SARS-CoV-2 infection—A comparative study from Novi Sad, Serbia. PLOS ONE. 2022 Feb 2;17:e0263468.

57. Müller L, Andrée M, Moskorz W, Drexler I, Walotka L, Grothmann R, et al. Age-dependent Immune Response to the Biontech/Pfizer BNT162b2 Coronavirus Disease 2019 Vaccination. Clin Infect Dis. 2021 Dec 6;73(11):2065–72.

58. Arvin AM, Fink K, Schmid MA, Cathcart A, Spreafico R, Havenar-Daughton C, et al. A perspective on potential antibody-dependent enhancement of SARS-CoV-2. Nature. 2020 Aug;584(7821):353–63.

59. Okuya K, Hattori T, Saito T, Takadate Y, Sasaki M, Furuyama W, et al. Multiple Routes of Antibody-Dependent Enhancement of SARS-CoV-2 Infection. Microbiology Spectrum. 2022 Mar 23;10(2):e01553–21.

60. Maemura T, Kuroda M, Armbrust T, Yamayoshi S, Halfmann PJ, Kawaoka Y. Antibody-Dependent Enhancement of SARS-CoV-2 Infection Is Mediated by the IgG Receptors FcγRIIA and FcγRIIIA but Does Not Contribute to Aberrant Cytokine Production by Macrophages. mBio. 2021 Oct 26;12(5):e0198721.

61. Ippoliti L, Coppeta L, Somma G, Bizzarro G, Borelli F, Crispino T, et al. Pulmonary function assessment after COVID-19 in vaccinated healthcare workers. J Occup Med Toxicol. 2023 Dec 15;18(1):31.

62. Vicenti I, Basso M, Gatti F, Scaggiante R, Boccuto A, Zago D, et al. Faster decay of neutralizing antibodies in never infected than previously infected healthcare workers three months after the second BNT162b2 mRNA COVID-19 vaccine dose. Int J Infect Dis. 2021 Nov;112:40–4.

63. Naaber P, Tserel L, Kangro K, Sepp E, Jürjenson V, Adamson A, et al. Dynamics of antibody response to BNT162b2 vaccine after six months: a longitudinal prospective study. The Lancet Regional Health - Europe. 2021 Sep 1;10:100208.

64. Ali H, Alahmad B, Al-Shammari AA, Alterki A, Hammad M, Cherian P, et al. Previous COVID-19 Infection and Antibody Levels After Vaccination. Front Public Health. 2021 Dec 1;9:778243.

65. Manisty C, Otter AD, Treibel TA, McKnight Á, Altmann DM, Brooks T, et al. Antibody response to first BNT162b2 dose in previously SARS-CoV-2-infected individuals. Lancet. 2021 Mar 20;397(10279):1057–8.

66. Bradley T, Grundberg E, Selvarangan R, LeMaster C, Fraley E, Banerjee D, et al. Antibody Responses after a Single Dose of SARS-CoV-2 mRNA Vaccine. N Engl J Med. 2021 May 20;384(20):1959–61.

67. Yamayoshi S, Yasuhara A, Ito M, Akasaka O, Nakamura M, Nakachi I, et al. Antibody titers against SARS-CoV-2 decline, but do not disappear for several months. EClinicalMedicine. 2021 Feb 11;32:100734.

68. Ferrari D, Ambrosi A, Di Resta C, Tomaiuolo R, Locatelli M, Banfi G. Evaluation of antibody titer kinetics and SARS-CoV-2 infections in a large cohort of healthcare professionals ten months after administration of the BNT162b2 vaccine. J Immunol Methods. 2022 Jul;506:113293.

